## Supplemental Table 1 for "Accessible LAMP-Enabled Rapid Test (ALERT) for detecting SARS-CoV-2"

Supplemental table 1: Primers

SARS-CoV-2 and Influenza B primers were found from the literature but modified for the sequence specific QUASR reporting system by us according to Ball et al. 2016. A gene with nasopharyngeal specific expression (Saferali et al. 2020), BPI fold containing family A, member 1 (BPIFA1), was used to design a sampling and process control using Primer Explorer v.4 (Eiken Chemical Co., Japan) to traverse exon/exon boundaries and therefore be mRNA specific (FIGURE). Primers were synthesized by IDT (Coraville, Iowa) and LGC Biosearch (Petaluma, CA). Primers modified with quenchers and TexasRed were purified by HPLC while all other primers were prepared by desalting. Lyophilized primers were reconstituted in water.

| **BPIFA1*** |  |
| --- | --- |
| FIP | ACAGCAGGCCATTGCTGAGGCTGCCCTTGAGTCCCACA |
| BIP | GAAAACCTTCCGCTCCTGGACATCCAAGCAGTCCCCCAAG |
| LF | GTCAAGCTTCCTGCAAGACC |
| LB-F | FAM-TCCTGAAGCCTGGAGGAGG |
| LB-Q | GGCTTCAGGA-BHQ1 |
| F3 | CCCTGCCCTTGAATGTGAAT |
| B3 | GGCCAGGAATCACTGACG |
| SARS-CoV-2 **NB/NA** | *Zhang et al. 2020* |
| FIP | TCTGGCCCAGTTCCTAGGTAGTTCGTGGTGGTGACGGTAA |
| BIP | AGACGGCATCATATGGGTTGCACGGGTGCCAATGTGATCT |
| LB | ACTGAGGGAGCCTTGAATACA |
| LF-TX** | 5TexRd-CCATCTTGGACTGAGATCTTTCATT |
| LF-Q | CAGTCCAAGATGG-3IAbRQSp |
| F3 | ACCGAAGAGCTACCAGACG |
| B3 | TGCAGCATTGTTAGCAGGAT |
| NA-F3 | TGGCTACTACCGAAGAGCT |
| **Influenza B** | *Ge et al. 2017* |
| FIP | TMARGGACAATACATTACGCATATCGATAAAGGAGGAAGTAAACACTCA |
| BIP | TAAAYGGAACATTCCTCAAACACCACTCTGGTCATAGGCATTC |
| LF | TYAAACGGAACTTCCCTTCTTTC |
| LB-F | FAM-GGATACAAGTCCTTATYAACTCTGC |
| LB-Q | AAGGACTTGTATTCC-BHQ1 |
| F3 | AGGGACATGAACAACAAAGA |
| B3 | CAAGTTTAGCAACAAGCCT |
| SARS-CoV-2 **NM** | *Broughton et al. 2020* |
| FIP-F | FAM-TGCGGCCAATGTTTGTAATCAGCCAAGGAAATTTTGGGGAC |
| FIP-Q | CATTGGCCGCA-IABkFQ |
| BIP | CGCATTGGCATGGAAGTCACTTTGATGGCACCTGTGTAG |
| LF | TTCCTTGTCTGATTAGTTC |
| LB | ACCTTCGGGAACGTGGTT |
| F3 | AACACAAGCTTTCGGCAG |
| B3 | GAAATTTGGATCTTTGTCATCC |

All sequences presented 5’ to 3’

Y represents degenerate C or T; R represents degenerate A or G; M represents degenerate A or C

* When used with NEB Colorimetric LAMP kit, a non-modified version of LB-F was used and LB-Q was omitted.

**In one iteration of this primer a FAM fluorophore is used at the 5’ end.
