## Supplemental Table 2 for "Accessible LAMP-Enabled Rapid Test (ALERT) for detecting SARS-CoV-2"

| Sample # | Known Value | Sample Type | Sample Source | Ct Value | Result | Result Interpretation | Notes |
|---|---|---|---|---|---|---|---|
| **Validation Trial Conducted In France** | | | | | | | |
| 1 | Negative | Cross Reactivity | Coronavirus NL63 | 0 | No Fluorescence | Negative | |
| 2 | Positive | Clinical | NP | 27.8 | Fluorescence | Positive | |
| 3 | Positive | Clinical | NP | 22.6 | Fluorescence | Positive | |
| 4 | Positive | Clinical | NP | 37.9 | Fluorescence | Positive | |
| 5 | Positive | Clinical | NP | 20.6 | Fluorescence | Positive | |
| 6 | Positive | Clinical | NP | 31 | No Fluorescence | Negative | False Negative |
| 7 | Positive | Clinical | NP | 28 | Fluorescence | Positive | |
| 8 | Positive | Clinical | NP | 23.6 | Fluorescence | Positive | |
| 9 | Positive | Clinical | NP | 18 | Fluorescence | Positive | |
| 10 | Positive | Clinical | NP | 34.3 | Fluorescence | Positive | |
| 11 | Positive | Clinical | NP | 23.6 | Fluorescence | Positive | |
| 12 | Positive | Clinical | NP | 19.6 | Fluorescence | Positive | |
| 13 | Positive | Clinical | NP | 19.1 | Fluorescence | Positive | |
| 14 | Positive | Clinical | NP | 36.1 | No Fluorescence | Negative | False Negative |
| 15 | Positive | Clinical | NP | 21.8 | Fluorescence | Positive | |
| 16 | Positive | Clinical | NP | 27 | No Fluorescence | Negative | False Negative |
| 17 | Positive | Clinical | NP | 24.5 | Fluorescence | Positive | |
| 18 | Positive | Clinical | NP | 35 | Fluorescence | Positive | |
| 19 | Positive | Clinical | NP | 13 | Fluorescence | Positive | |
| **Validation Trial Conducted In Chile** | | | | | | | |
| 1 | Negative | Clinical | NP | 0 | Faint Fluorescence | Possible positive - Retest | |
| 2 | Negative | Clinical | NP | 0 | No Fluorescence | Negative | |
| 3 | Negative | Clinical | NP | 0 | No Fluorescence | Negative | |
| 4 | Negative | Clinical | NP | 0 | No Fluorescence | Negative | |
| 5 | Negative | Clinical | NP | 0 | No Fluorescence | Negative | |
| 6 | Negative | Clinical | NP | 0 | No Fluorescence | Negative | |
| 7 | Negative | Clinical | NP | 0 | Faint Fluorescence | Possible positive - Retest | |
| 8 | Negative | Clinical | NP | 0 | Faint Fluorescence | Possible positive - Retest | |
| 9 | Negative | Clinical | NP | 0 | Faint Fluorescence | Possible positive - Retest | |
| 10 | Negative | Clinical | NP | 0 | No Fluorescence | Negative | |
| 11 | Positive | Clinical | NP | 31.68 | Fluorescence | Positive | |
| 12 | Positive | Clinical | NP | 34.57 | Fluorescence | Positive | |
| 13 | Positive | Clinical | NP | 35.34 | Faint Fluorescence | Possible positive - Retest | |
| 14 | Positive | Clinical | NP | 33.4 | Faint Fluorescence | Possible positive - Retest | |
| 15 | Positive | Clinical | NP | 34.32 | Fluorescence | Positive | |
| 16 | Positive | Clinical | NP | 32.2 | Fluorescence | Positive | |
| 17 | Positive | Clinical | NP | 32.34 | Fluorescence | Positive | |
| 18 | Positive | Clinical | NP | 34.17 | Faint Fluorescence | Possible positive - Retest | |
| 19 | Positive | Clinical | NP | 32.55 | Fluorescence | Positive | |
| 20 | Positive | Clinical | NP | 20.25 | Fluorescence | Positive | |
| 21 | Positive | Clinical | NP | 21.22 | Fluorescence | Positive | |
| 22 | Positive | Clinical | NP | 26.88 | Faint Fluorescence | Possible positive - Retest | |
| 23 | Positive | Clinical | NP | 26.11 | Fluorescence | Positive | |
| 24 | Positive | Clinical | NP | 24.16 | Fluorescence | Positive | |
| 25 | Positive | Clinical | NP | 27.68 | Fluorescence | Positive | |
| 26 | Positive | Clinical | NP | 29.29 | Faint Fluorescence | Possible positive - Retest | |
| 27 | Positive | Clinical | NP | 23.63 | Fluorescence | Positive | |
| 28 | Positive | Clinical | NP | 20.85 | Fluorescence | Positive | |
| 29 | Positive | Clinical | NP | 28.1 | Fluorescence | Positive | |

**Data from clinical validation conducted in France and Chile**
