## Supplemental Figure 1 for "Accessible LAMP-Enabled Rapid Test (ALERT) for detecting SARS-CoV-2"

a. b. c.

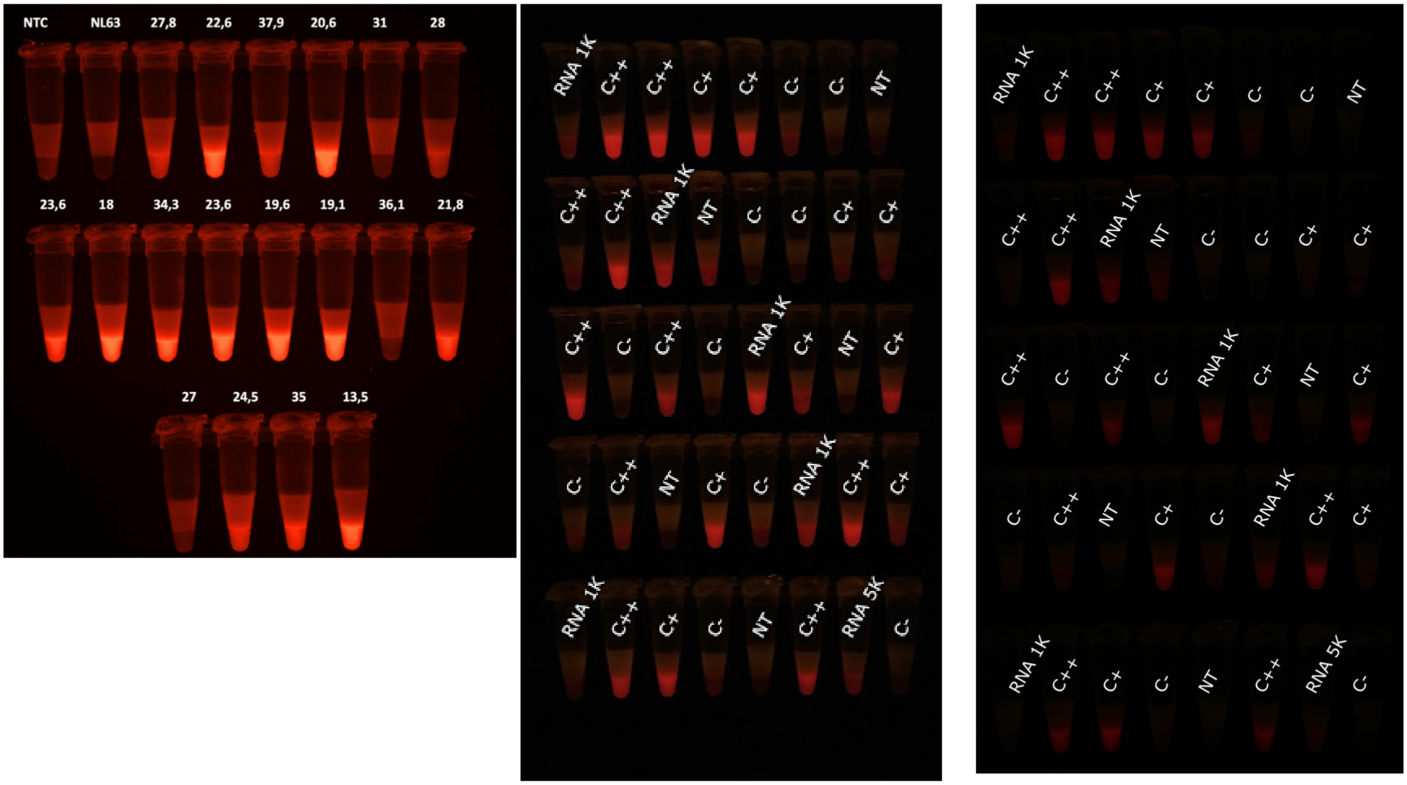


**A**. Validation study conducted at Hôpital Saint-Louis, Ct values are represented above detection cartridges, NL63 is cross-reactivity experiment with coronavirus NL63. **B** High exposure image of validation study conducted at Pontificia Universidad Católica de Chile, C++ represents samples with Ct below 30, C+ above 30 and C- above 40 or no amplification. RNA 1K and 5K represents positive control from in vitro transcription at 1000 or 5000 copies per reaction respectively **C.** Same tubes as B with a lower exposure image.
