## Supplemental Figure 2 for "Accessible LAMP-Enabled Rapid Test (ALERT) for detecting SARS-CoV-2"

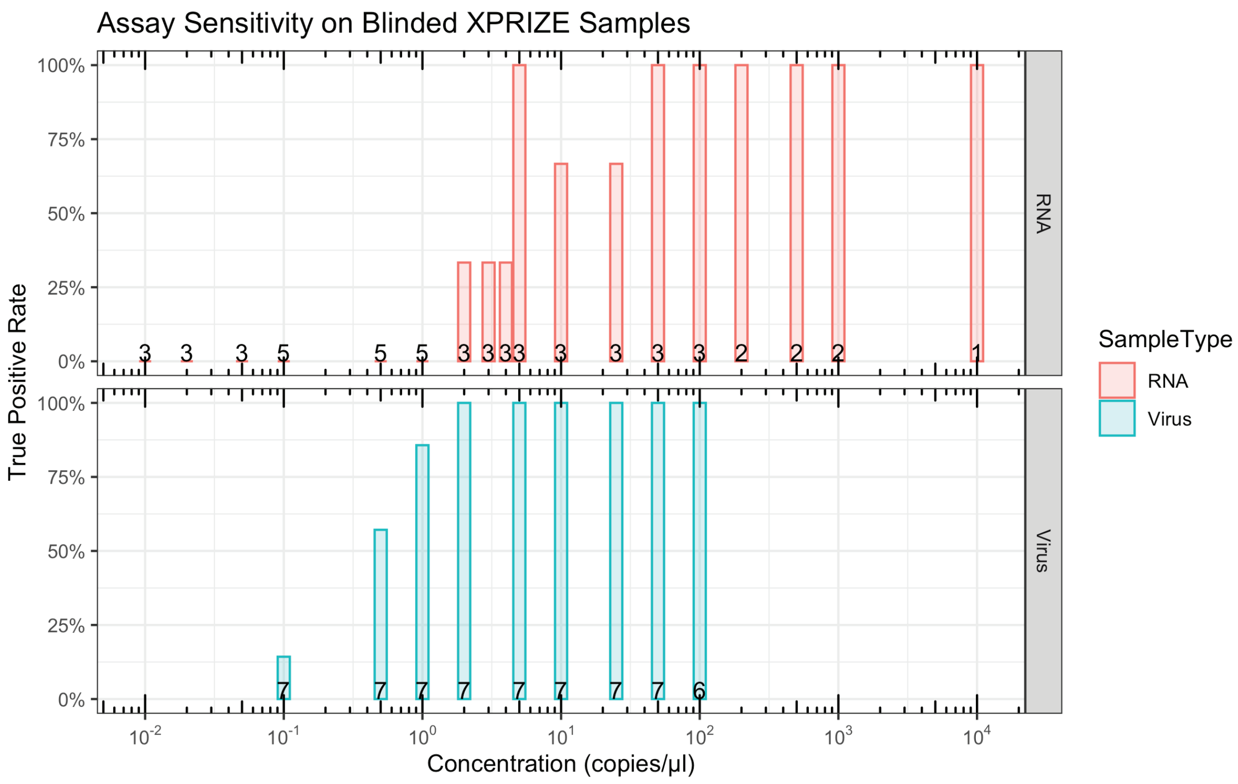


**Assay sensitivity on blinded XPRIZE samples for RNA vs virus**

True positive rate for our assay (y-axis) is shown for different concentrations (x-axis) for RNA vs virus. The number of samples tested at each concentration for a sample type is represented by the values above the x-axes.
